# Unveiling Circulating Targets in Pancreatic Cancer: Insights from Proteogenomic Evidence and Clinical Cohorts

**DOI:** 10.1101/2024.02.08.24302497

**Authors:** Haokang Feng, Zhixue Chen, Jianang Li, Jiale Feng, Fei Yang, Fansheng Meng, Hanlin Yin, Yuquan Guo, Huaxiang Xu, Yuxin Liu, Runjie Liu, Wenhui Lou, Liang Liu, Xu Han, Hua Su, Lei Zhang

## Abstract

Pancreatic cancer (PC), lacking biomarkers and effective therapeutics, remains highly lethal. Data regarding the correlations of PC risk and the individual plasma proteome known for minimally cancer biomarkers, are scarce. Here, we measure 1,345 human plasma proteins via Proteome-Wide Association Studies, presenting 78 proteins are prominently related to PC risk, including 4 proteins (ROR1, FN1, APOA5, ABO) exhibit the strongest causal association identified via Mendelian Randomization and Colocalization. Our two independent cohorts further demonstrate FN1 and ABO are highly expressed in blood or tumors from patients with PC compared to specimens from healthy individuals or para-tumors. Moreover, patients with higher levels of FN1 and ABO in their blood or tumors have worse median survival than those with lower levels. Multiple drugs targeting FN1 are currently available or undergoing clinical testing, making FN1 a promisingly repurposed therapeutic target in addition to severing as a circulating prognostic indicator for PC.

## Introduction

It remains a formidable challenge to management of pancreatic cancer (PC) whose overall survival (OS) rate is merely 12%.^1^ Most drugs targeting PC have been proven to be futile in clinical trials, with very limited efficacy in improving patient survival,^2^ while a few drugs only take effects in specific populations. A typical example is olaparib approved by the Food and Drug Administration (FDA) for germline BRCA-mutated (gBRCAm) metastatic PC.^3, 4^ Although olaparib exhibits a modest extension of progression-free survival (PFS), its efficacy is greatly compromised due to the low rate of BRCA mutation ranging from 4% to 7%.^5^ Thus, discovering more drug targets is urgent to improve treatment outcomes for diverse patients with PC.

Circulating blood, which flows through tumor sites and all organs, provides an extensive view of the pathophysiological state of individuals,^6^ and contributes to a more holistic understanding of the pathological and molecular genetic landscape of patients than does local pancreatic tumor sampling. Circulating proteins including functional proteins within the circulatory system and those leaked or secreted from tissues, have been widely used in identifying new diagnostic and therapeutic biomarkers for cancers,^7–10^ which requires to capture relevant low-abundance proteins from a vast pool of all circulating proteins.^11^ Combining high-throughput plasma proteomic sequencing with genetic association studies, enables the precise capture of potential targets within the vast pool. In 2022, the Atherosclerosis Risk in Communities (ARIC) study utilized the SOMAscan proteomics assay, successfully captured a total of 1,345 unique proteins from 9,084 participants, offering a comprehensive large-scale plasma protein spectrum.^12^ Leveraging popular genetic association research methods such as Protein-Wide Association Study (PWAS) and Mendelian Randomization (MR), it becomes reality to link genes to PC phenotypes through protein functional variations^13^ based on this extensive plasma protein spectrum. Moreover, MR is also able to use the random allocation of genetic variants to infer the genetically causal circulating proteins associated with PC.^14^ In contrast to previous small-scale observational studies, integrating of PWAS and MR resembles a natural large-scale randomized clinical trial, providing stronger evidential support and minimizing potential confounding factors.^15^ However, there is currently no existing research applying these approaches to identify potential drug targets for PC.

Here, we launched a systematically genetic association study. Employing PWAS and MR analyses, we narrowed down from 1,345 circulating proteins extracted from the ARIC study to four proteins (ROR1, FN1, APOA5, ABO) causally associated with PC. To validate the therapeutic potential of these proteins, we examined their expression levels in two independent single-center retrospective cohorts. While ABO and FN1 were highly expressed in blood and tumor tissues derived from patients with PC compared to samples collected from healthy participants and para-tumor tissues, there were no difference of expression levels of ROR1 and APOA5 among these blood samples. In addition, patients whose blood or tumors contained more ABO and FN1 had worse median survival than those had less of them. Notably, many drugs targeting FN1 and ROR1 are FDA approved or under clinical trials. Thus, we uncover causal association between circulating proteins and PC risk, and identify FN1 is a promising drug target and circulating prognostic indicator for PC.

## METHOD

### Human blood pQTL for Proteome-Wide Association Studies (PWAS)

We obtained 1,345 plasma protein data from the 7,213 European individuals-involved Atherosclerosis Risk in Communities (ARIC) study (http://nilanjanchatterjeelab.org/pwas/).^12^ The detailed information was found in the original research.

### Meta-GWAS for Pancreatic Cancer

We performed a meta-analysis using data from the UK Biobank (http://www.nealelab.is/uk-biobank)^16^ and the FinnGen consortium database (version R9, https://finngen.fi/en)^17^ to integrate genome-wide association study (GWAS) of PC. In total, 403 European patients with PC and 360,791 healthy participants from the UK Biobank, and 1,416 European patients and 287,137 healthy participants from FinnGen database were included in our study. The diagnosis of PC was determined based on the International Classification of Diseases codes (10th revision). The meta-analysis was applied with inverse-variance weighting and fixed effects using METAL software (version 2011-03-25, https://csg.sph.umich.edu/abecasis/Metal).^18^ After calculating the genomic inflation factor, we generated a Quantile-Quantile (QQ) Plot to compare the distribution of observed P-values with the expected distribution under the null hypothesis of no association (Fig. S1A), helping us avoid potential false positives. Additionally, we created a Manhattan plot to display the P-values for all genetic variants across the genome (R package qqman, https://github.com/stephenturner/qqman, Fig. S1B).^19, 20^

### PWAS

We systematically integrated the PC meta-GWAS with human blood proteomes obtained from the ARIC study. And PWAS was conducted with FUSION (https://github.com/gusevlab/fusion_twas).^21^ We utilized weights derived from the cis summary statistics of plasma proteins, selected based on significant non-zero cis-heritability estimated by GCTA (p < 0.01). Additionally, linkage disequilibrium (LD) reference weights tailored to individuals of EA from the 1000 Genomes Project (https://data.broadinstitute.org/alkesgroup/FUSION/LDREF.tar.bz2) were incorporated to enhance accuracy. The location of the genetic variants was transferred with LiftOver (http://genome.ucsc.edu/cgi-bin/hgLiftOver) across different version of human reference genome. The genetic impact of PC, represented by the PC GWAS z-score, was integrated with protein weights by computing a linear sum of z-score × weight for independent SNPs at the locus, and this procedure was performed with FUSION.

### Instrumental Variables (IVs) Selection

We further selected IVs for PWAS-significant proteins of PC using the summary statistics from a previous deCODE proteome study.^22^ IVs were chosen based on specific criteria: 1) Location restriction of cis-pQTL (variants within ±1 MB of the target protein coding genes boundaries); 2) Exclusion of variants within the Major Histocompatibility Complex (MHC) region (chr6 25.5–34.0 Mb); 3) Significant association with pQTL and serum protein levels (Pvalue ≤ 5 × 10^−8^); 4) Conducting LD clumping to identify independent pQTLs (r^2^ < 0.1); 5) Selection of SNPs with a strong association with the exposure, determined by an F statistic >10. The F statistics were computed using the formula: F = R^2^(N-K-1)/K(1-R^2^), with R^2^ calculated as R^2^ = 2 × MAF×(1-MAF) Beta^2^.^23^ The replication cohort for plasma protein pQTLs data was sourced from the extensive proteome INTERVAL study.^24^ This study encompassed the measurement of 2,994 plasma proteins in 3,301 European participants using the SomaLogic platform. Additionally, data from the KORA F4 study by Karsten Suhre et al.,^25^ which included 1,000 European participants and utilized the SOMAscan platform, was also incorporated for validation.

### Mendelian Randomization (MR)

In MR analysis, the exposure variable was employed with cis-pQTLs from the discovery cohorts, and the outcome as the results from PC meta-GWAS. MR analyses were conducted using the R package TwoSampleMR (https://mrcieu.github.io/TwoSampleMR/). We employed the inverse-variance weighted MR (IVW) method for instruments with more than one variant and the Wald-ratio method for single-variant instruments.^26^ Cochrane Q statistics were applied to assess the heterogeneity of genetic instruments,^27^ and MR-Egger was employed to identify potential horizontal pleiotropy,^28^ The significance threshold was set at Pvalue < 6.7×10^-4^ (0.05/74) after Bonferroni correction. In the replication cohort, we repeated the MR for the identified proteins or genes with a significance level of Pvalue < 0.05.

### Bayesian Colocalization Analysis

The colocalization analysis based on the R package coloc (https://github.com/chr1swallace/coloc) was conducted to assess the causality possibility of the disease associated variants across expression, protein and phenotypic level. The five hypotheses (H0-H4) were assigned as follow: p1=1×10^−4^, indicating the probability of one causal variant being exclusively associated with proteins; p2=1×10^−4^, indicating the likelihood of one causal variant having a sole impact on PC; and a joint prior probability, p12=5×10^−5^, representing the probability of one causal variant affecting both features. The posterior probability for H4 (PPH4) was established with a threshold of greater than 80% to denote a strong correlation. Furthermore, the additional sensitivity analysis on prior values was performed using the R package coloc,^29^ indicating that the specified values p12=5×10^−5^ yielded robust conclusions regardless of the scenarios (Fig. S2).

### Establishment of Single-Center Clinical Cohorts

To validate the possibility of identified actionable causal targets as biomarkers for PC, we established two cohorts. Both cohorts adhered to the principles of the Declaration of Helsinki and received approval from the Ethics Committees of Zhongshan Hospital, affiliated with Fudan University (B2023-342). Written informed consent was obtained from all patients before their inclusion.

The first cohort included 60 consecutive patients who underwent pancreaticoduodenectomy or distal pancreatectomy for PC and 100 healthy participants at Zhongshan Hospital between January 2022 and April 2022. All participants underwent fresh peripheral blood collection, and enzyme-linked immunosorbent assay (ELISA) analyses were performed to assess the expression levels of four blood proteins (ABO, FN1, APOA5, ROR1). The differences of protein expression between healthy individuals and patients were evaluated using two-tailed Mann-Whitney tests. The protein expression levels of the 60 patients were further categorized into high and low groups based on the median. To explore the association between the blood expression of these proteins and patient prognosis, PFS was defined as the outcome, defined as the duration from the first date of diagnosis to the date of disease progression or the last recorded follow-up examination. The relationship between protein expression levels and PFS was assessed using log-rank tests.

The second retrospective study involved an additional 96 consecutive patients with PC who had undergone pancreaticoduodenectomy at Zhongshan Hospital between March 2015 and December 2016. Tumor and adjacent tumor tissues collected during surgery were processed into tissue microarrays. Immunohistochemistry (IHC) was conducted to assess the expression levels of four druggable targets (ABO, FN1, APOA5, ROR1) between tumor and para-tumor tissues. Expression IHC scores were calculated by their expression level in dutal cells, fibroblasts or extracellular matrix (ECM), categorized into four groups: negative, weak, moderate, and strong. Paired t-tests were employed to compare IHC scores between tumor and para-tumor tissues. The low-expression group was defined as tumors with negative and weak score, while the high-expression group included those with moderate and strong score. The primary outcome was OS, defined as the duration from the date of diagnosis to the date of death or the last recorded follow-up examination.^30^ Log-rank tests were utilized to evaluate the relationship between expression levels and survival.

### Actionable Druggable Targets

To figure out whether any preexisting medications that were already approved or in clinical testing target the selected causative proteins, the ChEMBL dataset (version 32, https://www.ebi.ac.uk/chembl/),^31^ Drugbank dataset (version 5.1.10, https://go.drugbank.com/)^32^ and another publicly accessible data from OpenTargets (https://platform.opentargets.org/)^33^ were interrogated to provide the comprehensive information regarding their diseases diagnosis and treatment authorization status.

## RESULT

### Study design

Overview of the study design appears in Figure 1. We initially gathered GWAS summary statistics from the UK Biobank and the FinnGen consortium collaboration, which contain information about 403 European individuals with PC and 360,791 healthy participants from the UK Biobank, as well as 1,416 European patients and 287,137 healthy participants from the FinnGen database. Following that, we carried out a GWAS meta-analysis study using summary statistics from the two groups. Based on integrating 1,345 plasma proteins collected from the 7,213 participants-involved ARIC study with above meta-summary statistics, PWAS was then used to uncover possible PC-related proteins. After that, we used conditionally-independent genetic variants to carry out two-sample MR analysis to identify causal associated proteins of PC while addressing linkage disequilibrium by Bayesian Colocalization analysis. Finally, two independent single-center cohorts were set up to verify if the MR-identified causal proteins could be employed as biomarkers or drug targets in medical practice. In one cohort, we analyzed the levels of causal proteins in blood samples taken from 60 patients and 100 healthy volunteers, as well as the correlation of blood causal proteins from patients to PFS. In another cohort, we determined the relationship between the levels of the causal proteins and the prognosis by examining the levels of causal proteins in paired para-tumor and tumor specimens from 96 patients diagnosed with PC. Then the drug availability of the potential candidates was evaluated via CHEMBL dataset, DRUGBANK and OpenTargets.

**Figure. 1.**
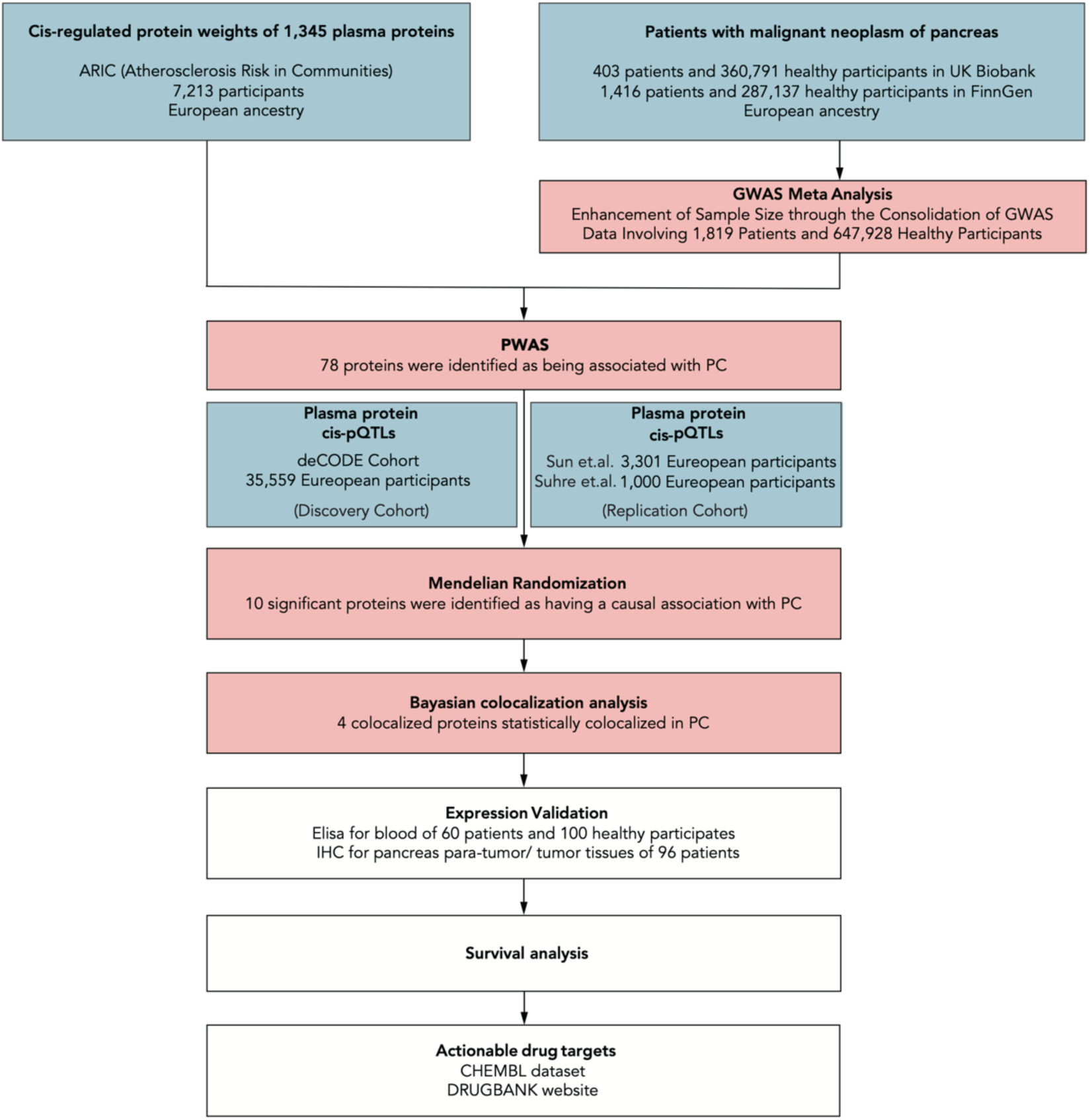
Overview of the study design

### PWAS discovers 78 circulating proteins correlated with PC

To uncover the correlated proteins with PC, we performed PWAS based on integrating the GWAS Meta Analysis and Elastic-net-based algorithms produced accurately predicted proteins in EA population from the ARIC study via using the FUSION pipeline.^21^ 78 circulating proteins were revealed to exhibit robust association with PC (Pvalue < 0.05) (Table S1, Fig. 2A). Further GO enrichment analysis on these 78 proteins identified their association with various biological processes, including collagen-containing extracellular matrix, sulfur compound metabolic, biosynthetic process and binding, vesicle lumen, secretory granule lumen, cytoplasmic vesicle lumen, sulfotransferase activity, heparin binding, transferase activity, transferring sulphur-containing groups, extracellular matrix binding, and collagen binding (Fig. 2B).

**Figure. 2.**
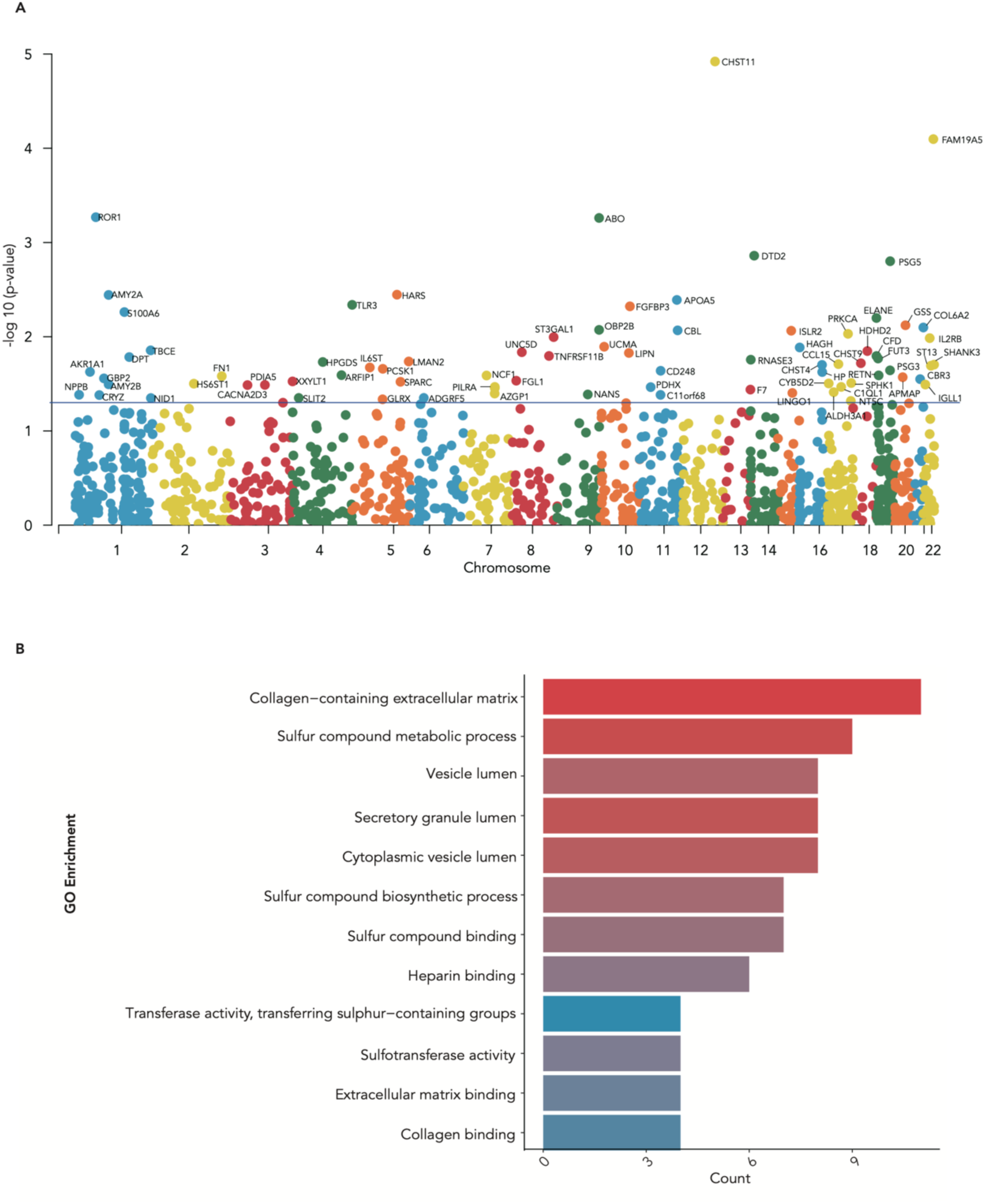
PWAS discovers 78 circulating proteins correlated with PC. (A) Manhattan plots illustrate PWAS analysis via integrating the PC meta-genome-wide association studies (GWAS) with the ARIC study. Each dot on the graph indicates a single association test result between a circulating target and PC. The x-axis represents the chromosomal location, and the y-axis shows -log10 (P-value). Dots below the blue line correspond to non-significant associations (Pvalue > 0.05), whereas dots above the line represent significant associations (Pvalue < 0.05). (B) GO analysis of 78 circulating proteins associated with PC identified by PWAS.

### 10 proteins are identified as suspected causal PC risk targets via MR analysis

To investigate potential causal associations between circulating proteins and PC, we conducted a systematic approach involving both MR and GWAS. Firstly, we used the deCODE proteomic data to identify conditionally independent cis-pQTLs for each circulating protein in the European cohort including 35,559 participants. A total of 4,002 cis-pQTLs were identified, covering 74 out of the 78 circulating proteins derived from above PWAS analysis. These cis-pQTLs demonstrated strong statistical power (F-statistic > 10). Next, we employed these cis-pQTLs as instrumental variables in a two-sample MR analysis using the meta-GWAS data for PC. This approach allowed us to identify 36 proteins with a significantly causal association with PC risk (Pvalue < 0.05). After applying Bonferroni correction (Pvalue< 0.05/74 = 6.7 × 10^-4^), only 11 proteins remained meaningful difference (Fig. 3, Table S2). We also assessed the robustness of our primary MR analyses by examining heterogeneity and horizontal pleiotropy. Our results indicated no observed heterogeneity (Q_pvalue > 0.05 in Table S2) and horizontal pleiotropy (Pvalue > 0.05) among the 10 causal proteins associated with PC. And only one protein OBP2B didn’t pass the heterogeneity (Q_pvalue=0.034) and horizontal pleiotropy (Pvalue = 0.013).

**Figure. 3.**
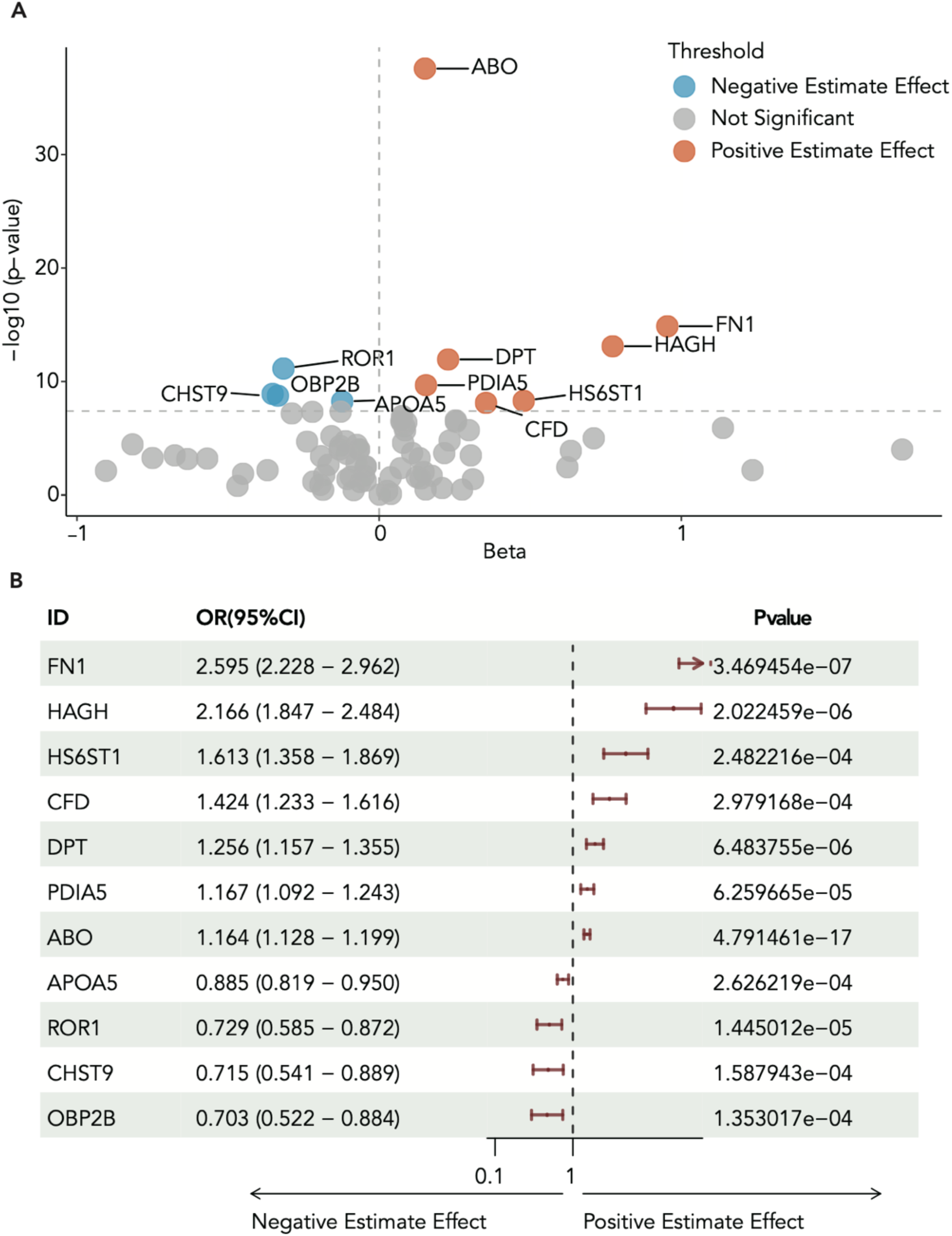
Mendelian Randomization (MR) for associations between circulating proteins and PC risk. (A) The genetic variants from the pQTL in the deCODE dataset were utilized for MR. The x-axis of volcano plot represents the beta value, while the y-axis indicates -log10 (Pvalue). Dots above the horizontal dashed line represent significant associations (Pvalue < 6.7×10-4 after Bonferroni correction). Red dots signify risk factors for PC, while blue dots represent protective factors for PC. (B) The forest plot showing the odds ratios and 95% confidence intervals of PC and 11 circulating targets after Bonferroni correction, however, OBP2B did not pass subsequent tests for pleiotropy and heterogeneity.

### PC risk proteins ABO, FN1 and ROR1 exhibit colocalization with pQTL

To assess whether a shared SNP influences both PC risk and protein expression levels, the MR-identified 10 protein targets were proceeded to colocalization analysis. Employing prior probabilities (p1=1×10^−4^, p2=1×10^−4^, p12=5×10^−5^) and posterior probabilities (PPH4>80%), we found that four (ROR1, FN1, APOA5, and ABO) out of the 10 proteins displayed strong genetic colocalization (PPH4 > 80%) (Fig. 4, Fig. S3). Utilizing data from the INTERVAL dataset and the KORA F4 study, we validaed the replicability (Pvalue < 0.05) of three plasma targets (ABO, FN1, and ROR1) when the IVW approach was implemented, but APOA5 failed to do so (Beta = -0.162, Pvalue = 0.244) (Table 1).

**Figure. 4.**
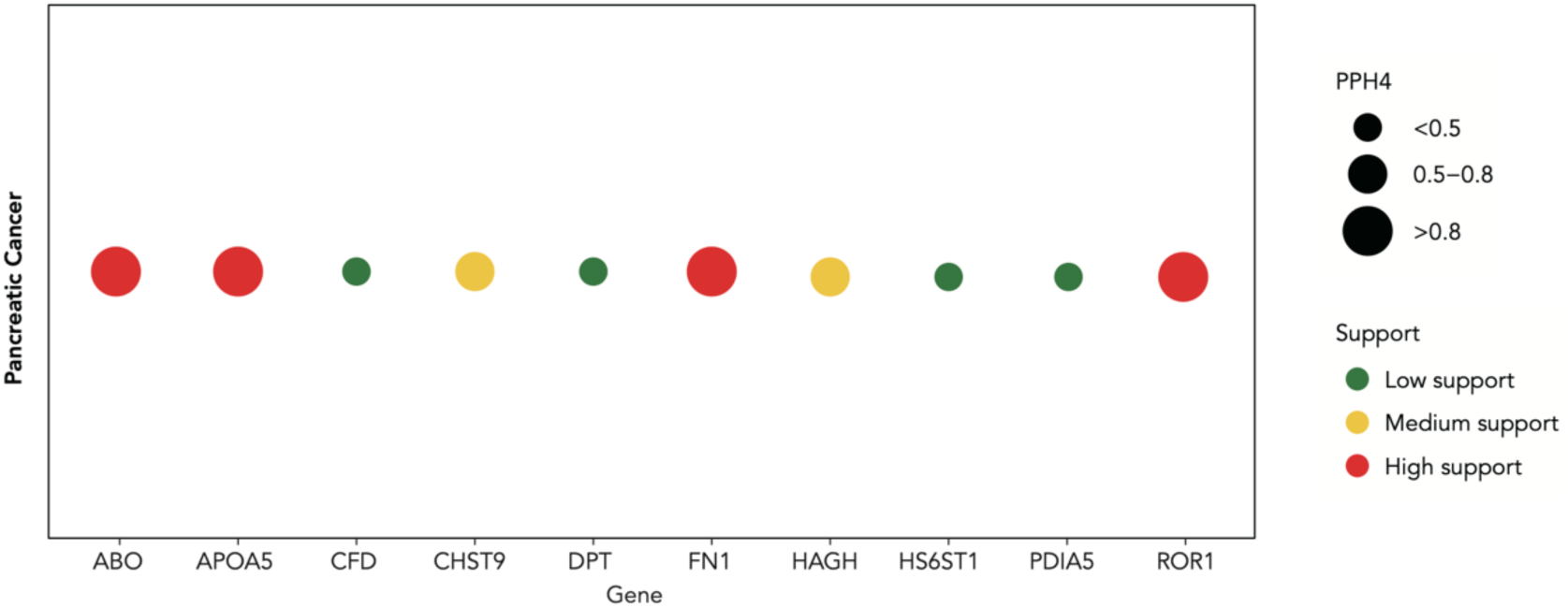
Regional association plots illustrating the colocalization analysis of four proteins causally associated with PC risk. The size of each circle corresponds to the colocalization P value for H4 (PPH4). When PPH4 >0.8, the two signals are considered to have strong evidence of colocalization. The color denotes the level of evidence, with red indicating high support.

**Table 1.**
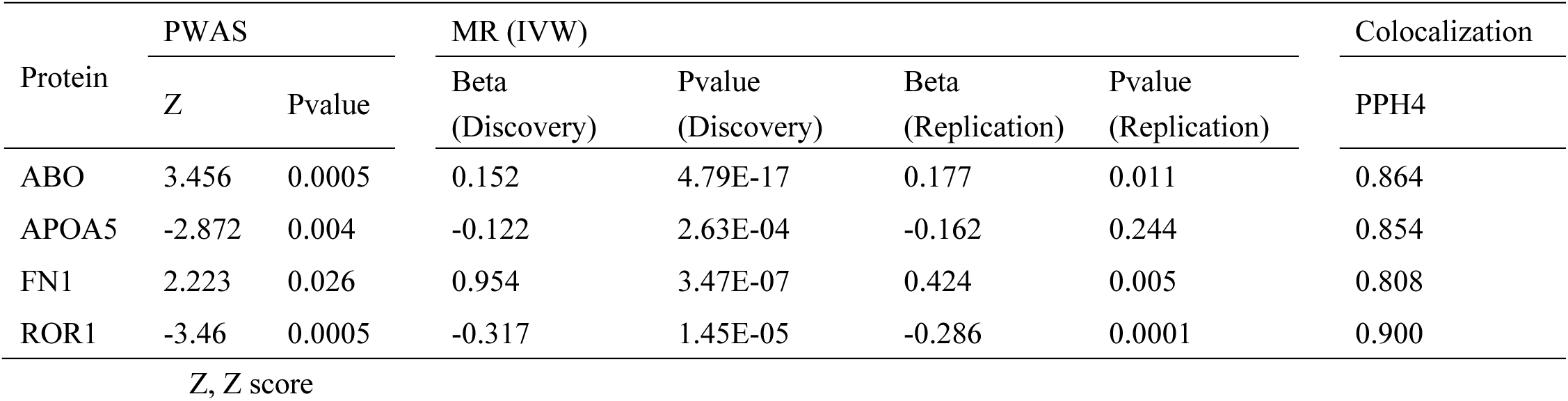
The MR results of four associated circulating targets in the discovery and replication cohorts.

### Elevated ABO and FN1 levels in peripheral blood and tumors are associated with reduced survival

In first retrospective clinical cohort, which included 60 patients with PC and 100 healthy individuals enrolled from January 2022 to April 2022, all patients underwent pancreaticoduodenectomy or distal pancreatectomy without any prior treatment. The baseline information was illustrated in Table S3. ELISA analysis of fresh peripheral blood samples collected from the individuls in this cohort revealed that ABO (Fig. 5A) and FN1 (Fig. 5B) proteins are highly expressed in patients with PC compared to healthy individuls, whereas no difference of ROR1 (Fig. 5C) and APOA5 (Fig. 5D) protein level is observed between these two groups. Upon dividing the expressions of the four proteins into their respective high and low groups according to the median values of all 60 patients, we found that patients with increased expression of ABO (Fig. 5E) or FN1 (Fig. 5F) experienced a shorter PFS than others. Conversely, the expression levels of ROR1 (Fig. 5G) and APOA5 (Fig. 5H) did not demonstrate any significant correlation with PFS in this cohort. The baseline character between two highly and lowly expressed groups was illustrated in Table S4. These results suggest ABO and FN1 are strongly circulating prognostic indicators.

**Figure. 5.**
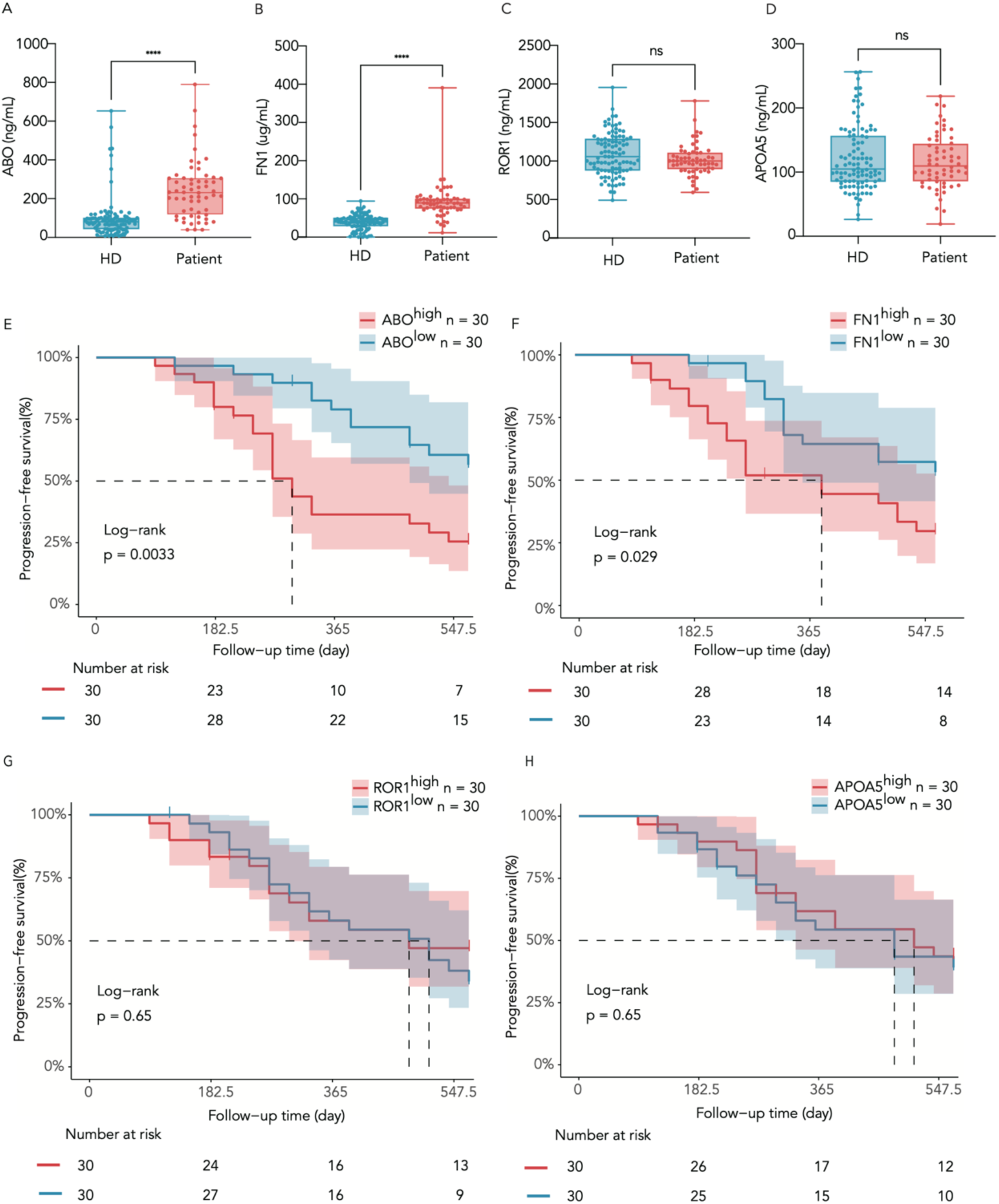
Blood expression levels of 4 causally associated proteins and their correlations with patient survival. ELISA analysis of plasma levels of (A) ABO, (B) FN1, (C) ROR1, and (D) APOA5 proteins between 100 healthy donors and 60 patients with PC. HD, healthy donors. (E-H) Progression Free Survival of patients with PC stratified according to median blood expression levels of above proteins. **** Pvalue < 0.0001, ns Pvalue > 0.05. Number at risk represents the number of individuals who have not experienced progression at the current time point.

In the second cohort consisting of 96 surgically resected human PC tumor and paired para-tumor tissues, spanning from March 2015 and December 2016. IHC analysis of these tissues revealed increased ABO (80/96), FN1 (85/96) and ROR1 (90/96) protein amounts in PC tumors compared to para-tumor tissues, whereas APOA5 (87/96) protein amounts are reduced in PC tumors relative to para-tumor tissues (Fig. 6A). Then we classified the expression levels of ABO, APOA5, FN1 and ROR1 in human PC tumors into 4 ranks, negative, weak, moderate and strong according to the IHC scores. This classification scheme defined negative and weak as the low-expression group, while moderate and strong were qualified as high-expression group (Fig. 6B). Propensity score matching (PSM) was performed to adjust the imbalanced baseline variables (Table S5). Patients with ABO^high^ or FN1^high^ tumors experienced worse median survival than those with low expression of these markers (Fig. 6C, D). However, no difference was observed between the APOA5 or ROR1 high and low groups (Fig. 6E, F). These findings are consistent with those obtained in first retrospective clinical cohort containg blood samples, suggesting that ABO and FN1 proteins are promising drug tagets in addition to seveing as prognostic biomarkers.

**Figure. 6.**
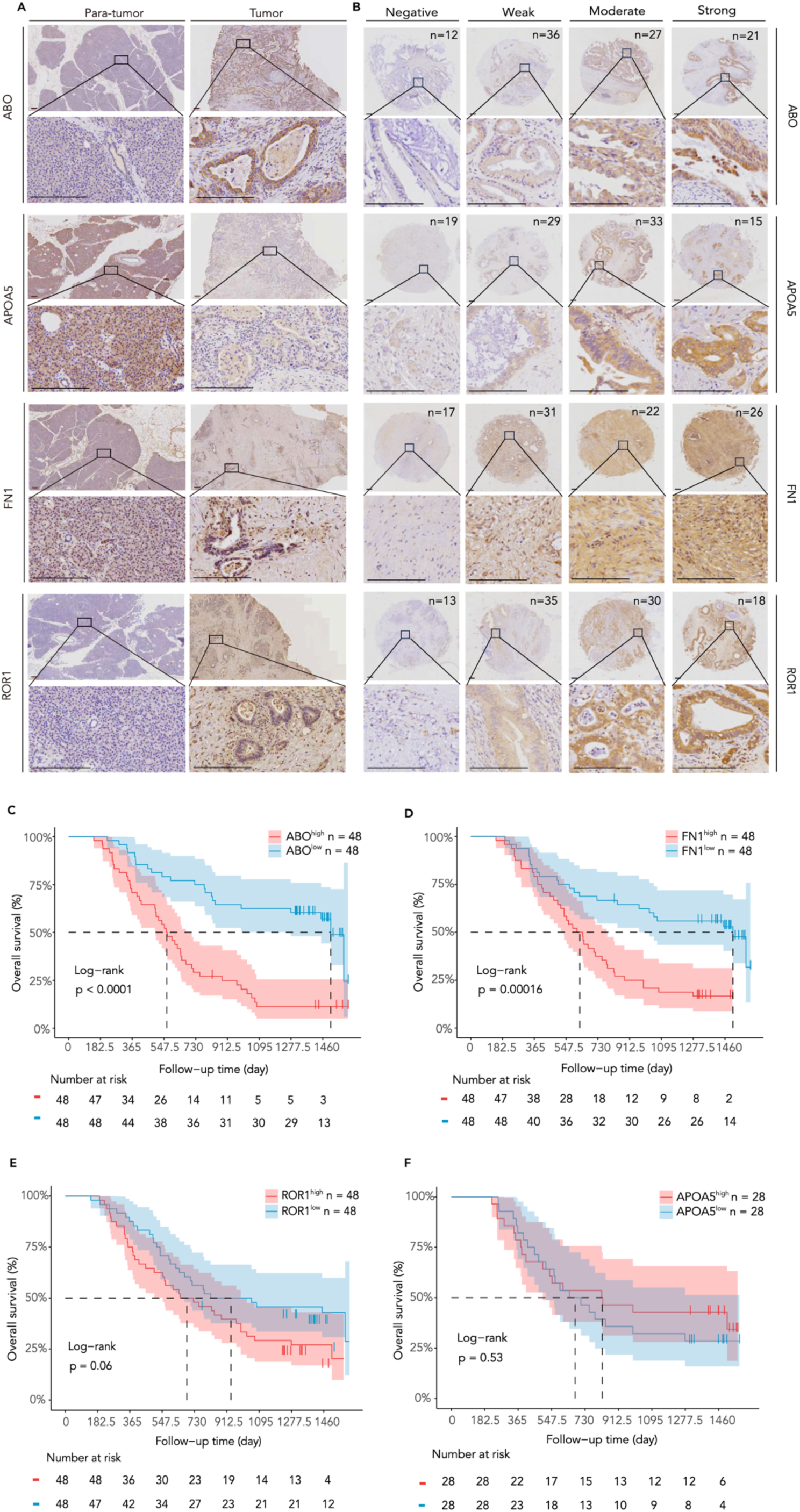
Tumor expression levels of 4 causally associated proteins and their correlations with patient survival. (A) Immunohistochemistry staining of ABO, FN1, APAO5, and ROR1 in paired PC tumor para-tumor tissues (n = 96). Scale bar, 150 μm. (B) PC tumors (n=96) were divided into four groups based on protein expression levels. Scale bar, 150 μm. The n on the top right corner of the upper figures represents the number of individuals at each level. (C-F) Overall survival of patients with PC stratified according to median tumor expression levels of the above four proteins. Number at risk represents the number of individuals still alive at the current time point.

### Identification of actionable Druggable Risk Targets

To explore the potential repurposed existing drugs for PC therapy, we investigated ABO, FN1, ROR1 and APOA5 as therapeutic targets by examing the ChEMBL, DrugBank, and OpenTargets datasets. Although no drugs on clincal trial were found to target ABO and APOA5, five compounds were found to target FN1, in which ocriplasmin has been approved by FDA for symptomatic vitreomacular adhesion (sVMA) therapy,^34^ and other compouds are currently undergoing clinical trials for various malignancies. In addition, the antibody cirmtuzumab against ROR1 is undergoing clinical trials for hematoma^35, 36^ and multiple solid neoplasms (NCT04504916). These results underscore the potential of FN1 and ROR1 as targets for drug repurposing in the treatment of PC (Fig. 7).

**Figure. 7.**
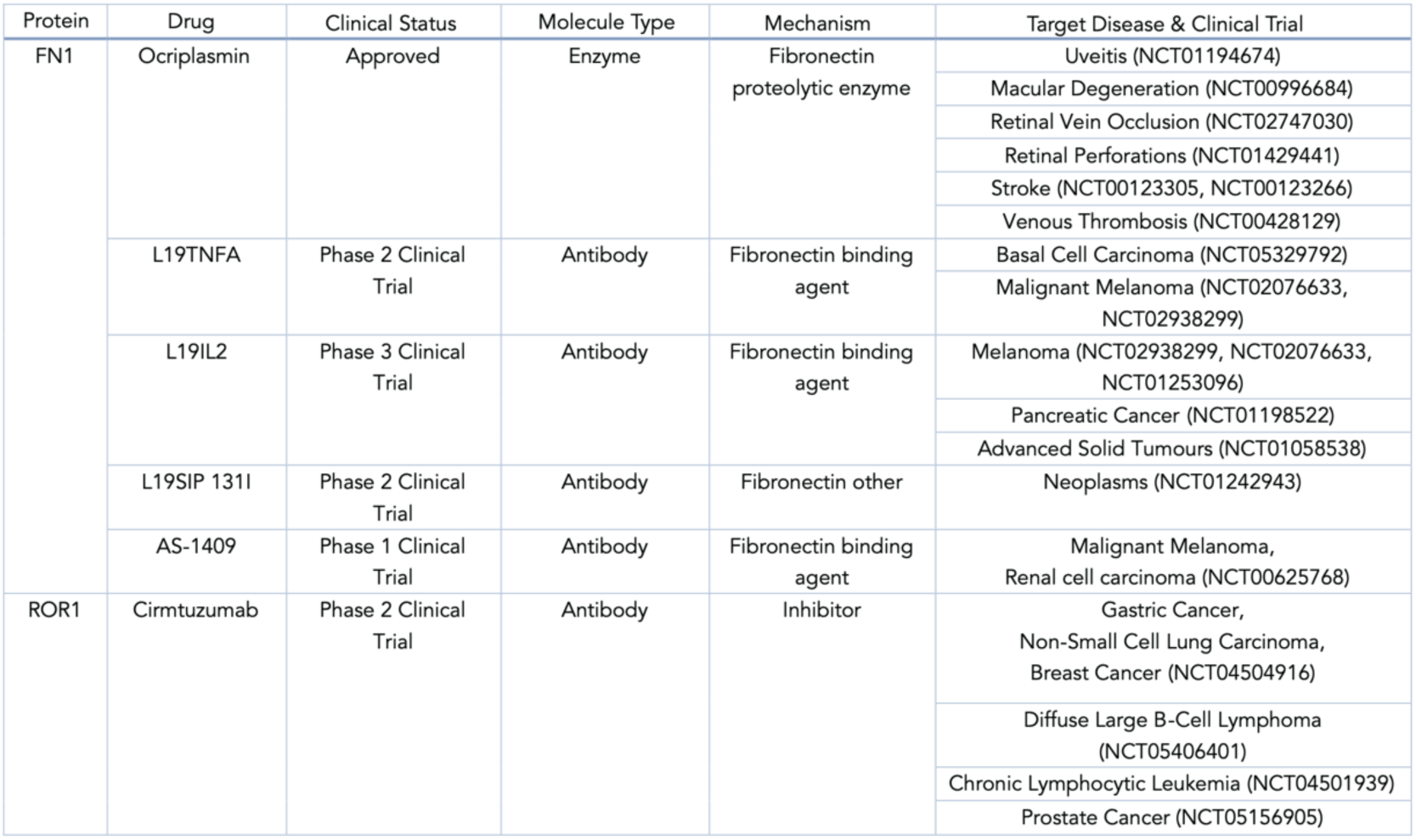
The actionable repurposing drugs of the causally associated targets.

### Discussion

Finding out additional biomarkers for diagnosis and drug targets is crucial for management of PC which displays highly heterogeneous among patients. Circulating proteomics characterized by its minimally invasive and global offers an attractive approach to identify functional biomarkers for PC. This study boasts a notable strength in its integration of vast population genetics information. Application of PWAS and MR allow us to explore the common genetic associations between circulating proteins and PC, offering robust evidence at both a high-throughput and genetic-causal level. In addition, we have successfully utilized two separate cohorts to confirm the presence of causal relationships between protein amounts and PC risk, providing valuable insights into their potential utility as biomarkers in monitoring PC.

Through applying GWAS-Meta analysis and ARIC study datasets, 78 circulating proteins were found to be associated with PC risk, which involved several secretary pathways, such as collagen-containing extracellular matrix, extracellular matrix binding, collagen binding and vesicle lumen, suggesting these circulating proteins may be secreted by tumor tissues. Four (ROR1, FN1, ABO, and APOA5) out of 78 proteins were shown the most causal association with PC identified via analyses of PWAS and MR. Although high expression of FN1 protein in PC tumors has been found to be associated with PC,^37–41^ it was previously unknown-to our knowledge-that FN1 protein levels in both blood and tumors of patients are strongly correlated with PC risk and patient survival. FN1, known as a widely expressed high-molecular-weight glycoprotein in the ECM,^42^ demonstrates high effectiveness in matrix metalloproteinase (MMP) production and close interaction with ECM proteins. The increased expression of FN1 in both blood and tumors and its relative to poor survivallend additionally support the concept that MMP-produced cleaved collagens contribute to the metabolic activity and progression of PC tumors.^30, 43–46^ In fact, collagen-III and MMP1 have been utilized for early detection of hepatocellular carcinoma,^47^ emphasizing the strong possibility that FN1could be used as a diagnostic and prognostic indicator for PC. Futhermore, the use of ocriplasmin, a drug already approved by the FDA to treat diabetic retinopathy,^48^ may be useful for treating PC, since it is an alpha-2 antiplasmin reducer and a truncated form of the natural human protein plasmin targeting FN1. By breaking down the protein matrix that cause mechanical compression, reducing interstitial fluid pressure, and improving drug absorption, ocriplasmin can potentially slow the growth of desmoplastic cancer, such as PC.

FN1 splice variant ED-B (ED-B) is a small domain composed of 91 amino acids, inserted into the FN1 molecule during the active tissue remodeling associated with angiogenesis which is critical to tumorigenesis,^49^ has been found to be accumulated around the new vascular systems in PC.^50, 51^ L19-IL2, an antibody against ED-B inhibited PC tumor growth and metastasis with remarable long-term tumor control in preclinical mouse models, greatly encouging to target FN1 to manage PC, while its safety has been validated in early clinical studies on patients with metastatic melanoma^52^ and advanced renal cell carcinoma.^53^ Indeed, a multicenter European Phase I study with an open-label, non-randomized, single-group assignment design (NCT01198522) further confirmed the safety and effectiveness of L19-IL2 combined with gemcitabine in patients with advanced PC, although the trial was terminated due to lack of recuitment.

The ABO gene encodes the glycosyltransferase enzyme responsible for major alleles (A and B), determining individuals’ blood types.^54^ Our findings suggest that tumor-secreted ABO may play a pivotal role in PC tumor progression, necessitating the identification of new glycosylated substrates of ABO. Variants in the ABO locus have also been linked to an increased risk of PC in individuals with non-O blood types,^55, 56^ and the relationship between ABO at both the genetic and transcriptional levels and susceptibility to PC has been established.^54, 57–59^ However, the causal connection between ABO proteins and PC had not been determined. Since proteins serve as direct executors of cellular functions, our results unequivocally prove the association of ABO to PC risk.

ROR1 is still a promising candidte for PC target based on its strong causative association to PC risk identified by MR analysis, and existing studies depiste of its expression levels being uncorrelated with pateints’ survival and lacking difference in the blood levels between healthy and patients with PC. Elevated levels of the circulating protein ROR1 in PC tumors compared to surrounding tissues are observed, which has been found to be associated with enhanced PC tumorigenic potential via the activation of AKT/c-Myc/E2F signaling pathways.^60^ Furthermore, the ROR1 Fz and Ig-like domain-developed humanized monoclonal antibody cirmtuzumab has shown encouraging safety and efficacy profiles in phase I/II clinical trials for hematological malignancies and breast cancer.^61–63^ Compared to the strong association of ROR1 and APOA5 with PC risk identified through PWAS and MR analyses in a large EA population, the relatively smaller size of the two independent cohorts may contribute to the disparity of ROR1 and APOA5 expression levels in blood and tumor specimens. Nonetheless, our findings reveal the causative circulating proteins of PC risk and strongly suggest that FN1 and ABO are promising drug targets for PC therapy.

### Limitations of the study

Our study is subject to several limitations. Firstly, the study utilized 1,345 instrumental variants from the ARIC study, but these variants do not encompass the entire range of actionable, druggable circulating proteins. Incorporating a wider spectrum of proteins could lead to the identification of more reliable therapeutic targets and blood biomarkers. Secondly, despite employing multiple data sources for a meta-analysis, the study was restricted to data from European individuals only. This limitation introduces the possibility of selection bias, underscoring the imperative for more expansive, multiethnic research initiatives to identify therapeutic targets that are specific to different populations. And this migh be one reason why the protein ROR1 and APOA5 could not be validated in our Asian clinical cohort. Thirdly, our study focused specifically on trans instrumental variables associated with pQTLs. Regrettably, this approach did not encompass the consideration of trans-acting regions and the intricate complexities of gene isoforms. Consequently, this may have implications for the precision of our causal estimates. Lastly, while we established two cohorts to assess the feasibility of identified targets as therapeutic candidates with matched biomarkers, it is essential to acknowledge that our sample size is limited and originates from a single center in Shanghai. To further validate the efficacy of these identified targets, additional researches involving larger-scale drug sensitivity tests, comprehensive animal experiments, and prospective studies are essential.

This study integrates multi-omics analyses and utilizes two single-center retrospective clinical cohorts to investigate the potential causal connections of PC. It furnishes compelling evidence across multiple levels to substantiate the role of circulating proteins as viable therapeutic targets for PC, accompanied by corresponding responsive biomarkers. Additionally, the study further discusses the potential molecular mechanisms associated with these molecules in PC. These findings contribute novel insights to the pharmaceutical management and pathogenesis of PC.

## RESOURCE AVAILABILITY

### Lead contact

Further information and requests should be directed to the lead contact, Hua Su.

### Materials availability

This study did not generate new unique reagents.

### Data and code availability data

GWAS summary statistics of the UK Biobank were obtained from http://www.nealelab.is/uk-biobank. GWAS summary statistics of the FinnGen (R9 release) were obtained from https://finngen.fi/en. The human blood pQTLs in PWAS was obtained from http://nilanjanchatterjeelab.org/pwas/. This paper does not report the original code.

## Acknowledgments

This work was supported by grants from the National Natural Science Foundation of China (82372884 to H.S., 82372644 and 82002931 to F.Y., 81972257, 82273382, 81827807, 81972218, 82103409, 82272929, 82173116), the Shanghai Natural Science Foundation (23ZR1413600 to H.S.), the Scientific Research StartingFoundation of Affiliated Zhongshan Hospital of Fudan University (2023ZSQD03 to H.S.); the Scientific Research Starting Foundation of Fudan University (JIH1340033Y to H.S.); the program of Qingfeng Schloarship of Shanghai Medical School of Fudan University (DGF819014/062 to H.S.), the Shanghai Higher Education Talent Program (SSF828065 to H.S.); Shanghai ShenKang Hospital Development Centre Project (SHDC2020CR2017B), Program of Shanghai Academic/Technology Research Leader (23XD1400600), China Postdoctoral Science Foundation (2021M690037), Shanghai Sailing Program (21YF1407100), Science and Technology Planning Project of Yunnan Province (202305AF150148), Shanghai Municipal Health Commission (20224Y0307), CSCO Cancer Research Fund Project (Y-HR2022QN-0085) and Open Fund of Key Laboratory of Hepatoaplenic Surgery, Ministry of Education, Harbin(GPKF202302). The funding agencies had no role in the study design, data collection and analyses, decision to publish, or preparation of the manuscript.

## Author contributions

H.F., H.S., L.Z., X.H. and L.L supervised and designed the study; H.F., Z.C., F.Y., F.M. and J.F. conducted the statistical analysis and experiments; H.S., H.F. and Z.C. drafted the manuscript; H.S., L.Z., X.H. and L.L. provided guidance and made revisions to the manuscript; H.Y., Y.G., H.X., Y.L., R.L. and W.L. collected experimental samples and clinical data. All authors contributed to and critically reviewed the manuscript.

