## Supplementary materials for "Unveiling Circulating Targets in Pancreatic Cancer: Insights from Proteogenomic Evidence and Clinical Cohorts"

Supplementary Appendix.

**Supplementary Fig. 1.** QQ plot and Manhattan plot of GWAS meta-analysis. (A) Deviations from the diagonal line observed in QQ plot demonstrated great association between SNP and genotype. The x-axis represents expected -log10 (Pvalue) and the y-axis represents the observed -log10 (Pvalue). (B) Manhattan plot demonstrated meta-analysis results for PC. The x-axis represents the chromosomal location, and the y-axis shows -log10 (Pvalue).

**Supplementary Fig. 2.** The post-hoc sensitivity analysis of the predefined prior probabilities. Sensitivity analysis revealed that for 10 circulating targets after MR, the predefined prior probability as p12=5×10^−5^ was all supported.

**Supplementary Fig. 3.** Regional association plots illustrating the colocalization analysis of 10 proteins with pancreatic cancer risk. The lead SNP is represented by a purple diamond. The threshold for p12 is set at 5×10^−5^, indicating the prior probability that a SNP is associated with both proteins and pancreatic cancer. Notably, the posterior probability (PPH4) values for ABO, APOA5, FN1, and ROR1 were all greater than 80%, providing strong evidence of colocalization.

**Supplementary Table 1.** 78 Circulating Proteins Robustly Associated with Pancreatic Cancer Identified by PWAS.

**Supplementary Table 2.** The MR, Pleiotropy and Heterogeneity Analysis of Eleven Associated Circulating Targets.

**Supplementary Table 3.** Characteristics of Patients with Pancreatic Cancer and Healthy Controls Undergoing ELISA.

**Supplementary Table 4.** Characteristics of Patients Stratified by Circulating Targets Expression Levels Evaluated by ELISA.

**Supplementary Table 5.** Characteristics of Patients Stratified by IHC Score Levels.

This supplemental material has been provided by the authors to give readers additional information about their work.

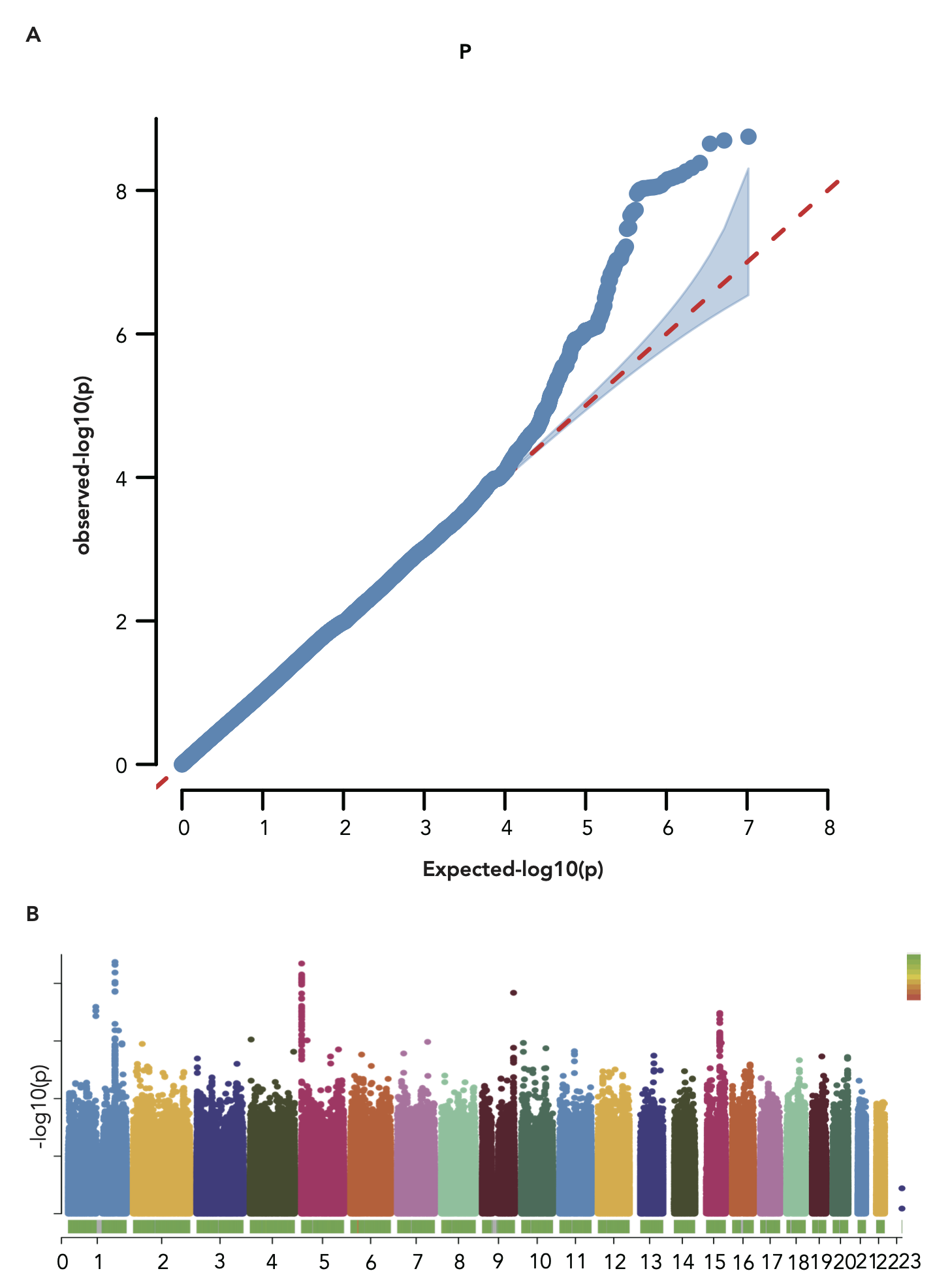

Figure S1. QQ plot and Manhattan plot of GWAS meta-analysis. (A) Deviations from the diagonal line observed in QQ plot demonstrated great association between SNP and genotype. The x-axis represents expected -log10 (Pvalue) and the y-axis represents the observed -log10 (Pvalue). (B) Manhattan plot demonstrated meta-analysis results for PC. The x-axis represents the chromosomal location, and the y-axis shows -log10 (Pvalue).

Figure S2. The post-hoc sensitivity analysis of the predefined prior probabilities. Sensitivity analysis revealed that for 10 circulating targets after MR, the predefined prior probability as p12=5×10^−5^ was all supported.

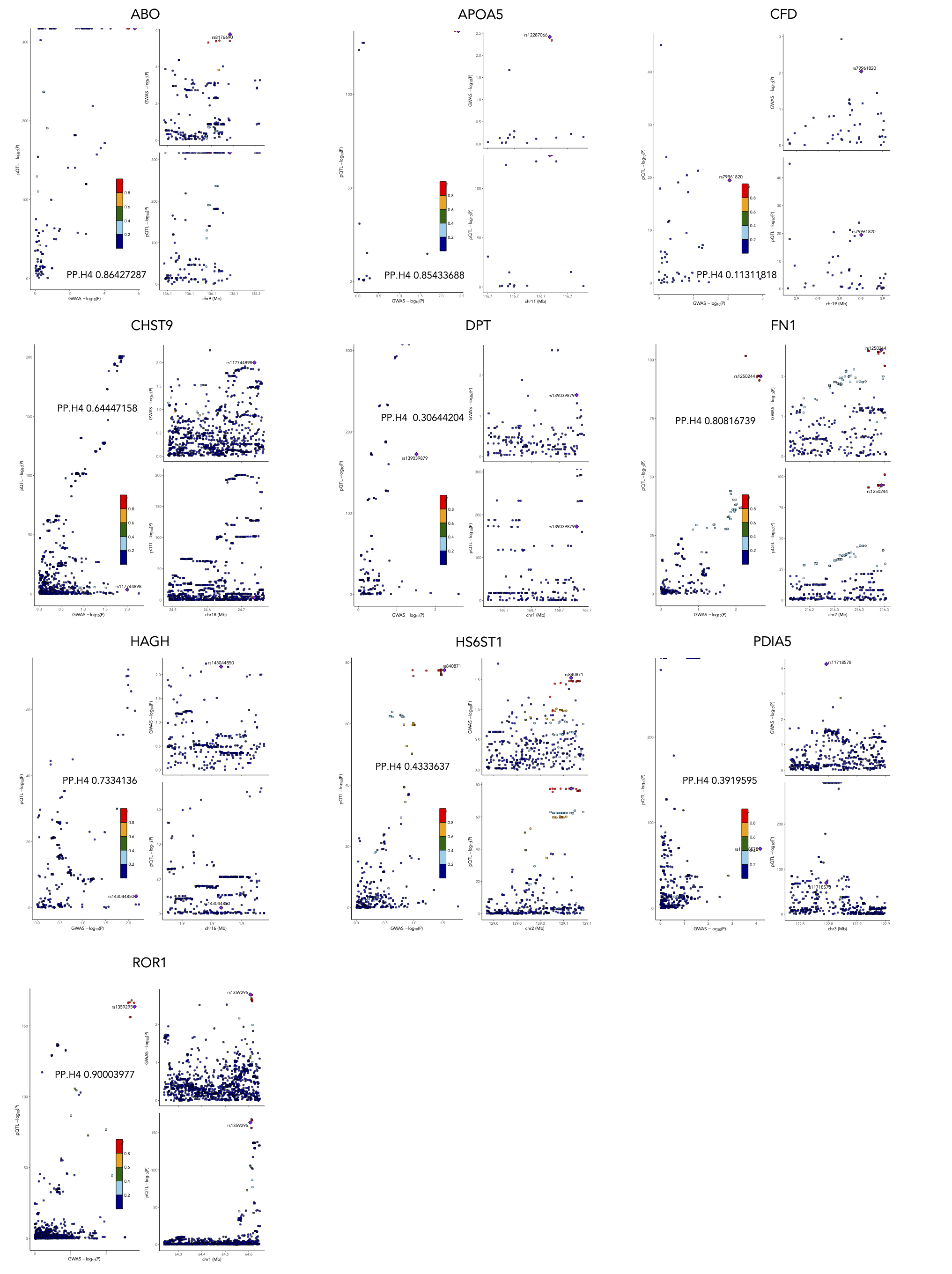

Figure S3. Regional association plots illustrating the colocalization analysis of 10 proteins with pancreatic cancer risk. The lead SNP is represented by a purple diamond. The threshold for p12 is set at 5×10^−5^, indicating the prior probability that a SNP is associated with both proteins and pancreatic cancer. Notably, the posterior probability (PPH4) values for ABO, APOA5, FN1, and ROR1 were all greater than 80%, providing strong evidence of colocalization.

**Table S1.** **78 Circulating Proteins Robustly Associated with Pancreatic Cancer Identified by PWAS.**

| **ID** | **Chr** | **Best** **GWAS ID** | **Best GWAS** **Z** | **pQTL ID** | **pQTL Z** | **nSNP** | **PWAS** **Z** | **PWAS Pvalue** |
| --- | --- | --- | --- | --- | --- | --- | --- | --- |
| CHST11 | 12 | rs1650127 | -3.973 | rs1704878 | -31.38 | 113 | 4.3774 | 0.000012 |
| FAM19A5 | 22 | rs5768649 | -2.98 | rs5768649 | -6.42 | 33 | 3.944494 | 0.00008 |
| ROR1 | 1 | rs6686041 | 3.14 | rs2224876 | -29.64 | 134 | -3.4605 | 0.000539 |
| ABO | 9 | rs10901250 | 3.373 | rs8176632 | 45.9 | 34 | 3.4555 | 0.000549 |
| DTD2 | 14 | rs17097892 | -2.6 | rs17097898 | -18.44 | 67 | 3.19917 | 0.00138 |
| PSG5 | 19 | rs150744069 | -3.579 | rs7253874 | -35.74 | 115 | -3.1591 | 0.00158 |
| HARS | 5 | rs116774392 | -3.303 | rs112352450 | 7.68 | 22 | -2.9132 | 0.00358 |
| AMY2A | 1 | rs72690677 | 2.907 | rs12076610 | 24.09 | 143 | 2.9109 | 0.003604 |
| APOA5 | 11 | rs60972755 | -2.977 | rs35120633 | 39.14 | 17 | -2.87272 | 0.00407 |
| TLR3 | 4 | rs36035924 | -2.76 | rs3775291 | -42.95 | 73 | 2.8343 | 0.00459 |
| FGFBP3 | 10 | rs7095552 | -2.88 | rs11186737 | -33.33 | 52 | 2.822918 | 0.00476 |
| S100A6 | 1 | rs2265081 | -2.294 | rs60969679 | 12.14 | 24 | -2.7778 | 0.005474 |
| ELANE | 19 | rs58350690 | -3.185 | rs56283881 | -12.58 | 41 | 2.7299 | 0.00633 |
| GSS | 20 | rs78882575 | 2.93 | rs6088642 | 25.78 | 36 | -2.671 | 0.00756 |
| COL6A2 | 21 | rs1980983 | 3.258 | rs35548026 | -18.56 | 73 | 2.6528 | 0.00798 |
| OBP2B | 9 | rs4962103 | -2.684 | rs4454354 | 17.3 | 20 | -2.6331 | 0.00846 |
| CBL | 11 | rs113715244 | -3.205 | rs76683040 | -21.75 | 89 | 2.62899 | 0.00856 |
| ISLR2 | 15 | rs62005678 | 3.02 | rs11632698 | -16.37 | 64 | 2.62614 | 0.00864 |
| PRKCA | 17 | rs61762372 | -2.343 | rs61762372 | 17.84 | 80 | -2.6 | 0.00932 |
| ST3GAL1 | 8 | rs11575971 | -2.659 | rs9643300 | -21.08 | 65 | -2.5724 | 0.0101 |
| IL2RB | 22 | rs228953 | -2.26 | rs228953 | 9.26 | 20 | -2.563922 | 0.01035 |
| UCMA | 10 | rs2093847 | 2.89 | rs1537771 | -18.67 | 96 | -2.490153 | 0.01277 |
| HAGH | 16 | rs2294620 | 2.616 | rs116869551 | 23.32 | 77 | 2.4831 | 0.013 |
| TBCE | 1 | rs10802904 | 2.836 | rs4469707 | -23.07 | 89 | 2.4584 | 0.013958 |
| HDHD2 | 18 | rs75284103 | -2.52 | rs79231363 | 36.68 | 106 | 2.452 | 0.0142 |
| UNC5D | 8 | rs189879235 | -3.161 | rs3108622 | -17.3 | 59 | -2.441 | 0.0146 |
| LIPN | 10 | rs10887850 | -2.57 | rs10509554 | 44.7 | 25 | 2.435592 | 0.01487 |
| TNFRSF11B | 8 | rs4629902 | 2.283 | rs2468184 | 8.38 | 92 | 2.4089 | 0.016 |
| CFD | 19 | rs74609243 | 3.586 | rs72984031 | -10.38 | 53 | 2.4085 | 0.01602 |
| DPT | 1 | rs80041579 | 2.284 | rs1018454 | 25.7 | 121 | 2.399 | 0.016441 |
| FUT3 | 19 | rs892140 | -3.11 | rs708686 | -37.04 | 116 | -2.3858 | 0.01704 |
| RNASE3 | 14 | rs11626000 | 2.44 | rs147307766 | 24.79 | 181 | -2.37549 | 0.01753 |
| LMAN2 | 5 | rs2029164 | -2.679 | rs35582636 | 6.65 | 61 | -2.3593 | 0.01831 |
| HPGDS | 4 | rs6840479 | -2.13 | rs10033662 | 33.27 | 63 | -2.3544 | 0.01855 |
| CHST9 | 18 | rs2114471 | -2.51 | rs11660451 | 17 | 41 | -2.343 | 0.0191 |
| CCL15 | 17 | rs2293788 | 2.739 | rs854624 | 45.51 | 32 | -2.3353 | 0.01953 |
| SHANK3 | 22 | rs151247655 | -2.36 | rs6009946 | 21.34 | 25 | -2.330492 | 0.01978 |
| CHST4 | 16 | rs75768020 | 2.437 | rs16973264 | 11.22 | 27 | 2.3284 | 0.0199 |
| ST13 | 22 | rs138337 | 2.39 | rs138337 | 16.86 | 21 | 2.320353 | 0.02032 |
| IL6ST | 5 | rs6894414 | -2.281 | rs13183065 | 25.42 | 61 | -2.3045 | 0.0212 |
| PCSK1 | 5 | rs13169290 | -2.368 | rs13169290 | -53.66 | 43 | 2.2918 | 0.02191 |
| PSG3 | 19 | rs10417319 | -4.124 | rs2355433 | -43.28 | 108 | -2.2776 | 0.02275 |
| CD248 | 11 | rs490972 | -2.771 | rs565972 | 5.71 | 18 | -2.27383 | 0.02298 |
| AKR1A1 | 1 | rs75598025 | 2.956 | rs2229540 | -40.86 | 51 | -2.2634 | 0.023612 |
| HP | 16 | rs2336601 | 2.805 | rs11648003 | -26.07 | 49 | -2.2626 | 0.0237 |
| ARFIP1 | 4 | rs11099853 | 2.49 | rs4619875 | 25.07 | 28 | 2.2326 | 0.02557 |
| RETN | 19 | rs73493937 | 2.044 | rs34124816 | -14.93 | 112 | -2.2304 | 0.02572 |
| NCF1 | 7 | rs148581667 | 2.899 | rs148581667 | 20.11 | 128 | 2.2295 | 0.0258 |
| FN1 | 2 | rs1250258 | 2.78 | rs1250258 | 15.11 | 24 | 2.2229 | 0.0262 |
| APMAP | 20 | rs4539860 | 2.63 | rs6036977 | -8.88 | 21 | -2.2132 | 0.02688 |
| GBP2 | 1 | rs2031834 | 1.951 | rs10922556 | 7.97 | 25 | 2.2018 | 0.027679 |
| CBR3 | 21 | rs79885774 | 2.906 | rs60376898 | -48.63 | 74 | -2.1947 | 0.02819 |
| FGL1 | 8 | rs7838153 | 2.877 | rs3739406 | 40.11 | 51 | 2.1789 | 0.0293 |
| XXYLT1 | 3 | rs752206 | -1.98 | rs13072883 | 17.83 | 57 | -2.1729 | 0.0298 |
| SPARC | 5 | rs115362461 | -2.386 | rs59311424 | -8.05 | 32 | 2.1691 | 0.03008 |
| SPHK1 | 17 | rs383048 | 2.554 | rs12941069 | 8.77 | 76 | -2.1572 | 0.03099 |
| CYB5D2 | 17 | rs72829384 | 1.751 | rs183282334 | -19.13 | 34 | 2.153 | 0.03132 |
| HS6ST1 | 2 | rs4662790 | -2.091 | rs4662790 | -14.99 | 99 | 2.15026 | 0.0315 |
| AMY2B | 1 | rs115608211 | 2.519 | rs12076610 | 21.42 | 145 | 2.1446 | 0.031985 |
| IGLL1 | 22 | rs187060998 | 2.76 | rs9624216 | -17.62 | 64 | -2.141737 | 0.03221 |
| PDIA5 | 3 | rs137867617 | 2.48 | rs3804749 | -58.98 | 102 | 2.1378 | 0.0325 |
| CACNA2D3 | 3 | rs62255460 | -2.32 | rs6802227 | 16.36 | 114 | 2.1366 | 0.0326 |
| C1QL1 | 17 | rs3744477 | -3.128 | rs7225162 | -19.9 | 56 | -2.1192 | 0.03408 |
| PILRA | 7 | rs2405442 | -2.24 | rs1859788 | -61.01 | 11 | 2.1183 | 0.0341 |
| PDHX | 11 | rs10836375 | -3.518 | rs2915221 | -18.83 | 143 | -2.11682 | 0.03427 |
| F7 | 13 | rs555212 | 2.45 | rs6046 | -48.3 | 77 | -2.0919 | 0.0364 |
| ALDH3A1 | 17 | rs2108967 | 2.653 | rs887241 | 12.46 | 12 | 2.0669 | 0.03875 |
| LINGO1 | 15 | rs4886869 | -3.57 | rs62007781 | 7.99 | 68 | -2.05867 | 0.03953 |
| AZGP1 | 7 | rs117268080 | -2.538 | rs1981550 | -28.56 | 21 | -2.0524 | 0.0401 |
| NANS | 9 | rs72759614 | -2.733 | rs7855984 | 10.44 | 95 | -2.0435 | 0.041005 |
| NPPB | 1 | rs41394446 | 2.554 | rs198379 | 12.01 | 25 | -2.0403 | 0.04132 |
| C11orf68 | 11 | rs490972 | -2.771 | rs554169857 | 15.05 | 26 | -2.03942 | 0.04141 |
| CRYZ | 1 | rs4650280 | 2.284 | rs3819946 | 44.63 | 52 | 2.0375 | 0.041599 |
| ADGRF5 | 6 | rs60401949 | 2.74 | rs586024 | 40.97 | 59 | -2.0086 | 0.0446 |
| SLIT2 | 4 | rs74377837 | 2.85 | rs587668 | -8.57 | 65 | -2.008 | 0.04465 |
| NID1 | 1 | rs56275761 | -2.162 | rs2734807 | 13.42 | 70 | -2.0069 | 0.044759 |
| GLRX | 5 | rs10476544 | 2.254 | rs6556884 | -6.33 | 17 | -1.994 | 0.04615 |
| NT5C | 17 | rs117008429 | 3.251 | rs78625720 | -29.24 | 67 | -1.9772 | 0.04802 |

PWAS, Proteome-Wide Association Study; Chr, chromosome; GWAS, Genome Wide Association Study; Z, Z score; pQTL, protein quantitative trait loci; SNP, Single Nucleotide Polymorphisms

**Table S2.** The MR, Pleiotropy and Heterogeneity Analysis of Eleven Associated Circulating Targets.

| Protein | MR (IVW) | | |  | Pleiotropy | |  | Heterogeneity (Cochran's Q Test) | |
| --- | --- | --- | --- | --- | --- | --- | --- | --- | --- |
|  | Beta | SE | Pvalue |  | MR-Egger-intercept | Pvalue |  | MR-Egger. Q | Q_pvalue |
| ABO | 0.152 | 0.018 | 4.79E-17 |  | 0.008 | 0.258 |  | 350.140 | 0.062 |
| ROR1 | -0.317 | 0.073 | 1.45E-05 |  | 0.02 | 0.367 |  | 37.356 | 0.633 |
| FN1 | 0.954 | 0.187 | 3.47E-07 |  | 0.008 | 0.845 |  | 4.284 | 0.891 |
| HAGH | 0.773 | 0.163 | 2.02E-06 |  | 0.006 | 0.887 |  | 3.587 | 0.964 |
| APOA5 | -0.122 | 0.034 | 2.60E-4 |  | -0.002 | 0.831 |  | 99.664 | 0.406 |
| DPT | 0.228 | 0.051 | 6.48E-06 |  | -0.015 | 0.231 |  | 78.985 | 0.634 |
| CHST9 | -0.335 | 0.089 | 1.59E-4 |  | 0.01 | 0.708 |  | 22.990 | 0.879 |
| HS6ST1 | 0.478 | 0.130 | 2.48E-4 |  | 0.051 | 0.086 |  | 8.424 | 0.935 |
| PDIA5 | 0.155 | 0.039 | 6.26E-05 |  | 0.007 | 0.567 |  | 107.832 | 0.620 |
| OBP2B | -0.353 | 0.092 | 1.35E-4 |  | -0.054 | 0.013 |  | 58.993 | 0.034 |
| CFD | 0.354 | 0.098 | 2.98 E-4 |  | 0.014 | 0.576 |  | 30.783 | 0.626 |

MR, mendelian randomization; IVW, inverse variance weighted method; SE, standard error; Pvalue, P value.

**Table S3. Characteristics of Patients with Pancreatic Cancer and Healthy Controls Undergoing ELISA.**

| Characteristics | Non-tumor  N=100 | Tumor  N=60 | Pvalue |
| --- | --- | --- | --- |
| Demographics and history |  | |  |
| Age, median (IQR) | 52 (18.00) | 66 (12.00) | **<0.001** |
| Male, n (%) | 49 (49.0%) | 40 (66.7%) | **0.029** |
| BMI>28 (kg/m^2^), n (%) | 13 (3.0%) | 9 (15.0%) | 0.722 |
| Hypertension, n (%) | 30 (30.0%) | 26 (43.4%) | 0.087 |
| Diabetes, n (%) | 40 (40.0%) | 32 (53.3%) | 0.101 |
| CVD, n (%) | 19 (19.0%) | 19 (31.0%) | 0.068 |
| Stroke, n (%) | 2 (2.0%) | 3 (5.0%) | 0.364 |
| Non-O blood type, n (%) | 71 (71.0%) | 46 (76.7%) | 0.434 |

BMI, Body Mass Index; CVD, coronary heart disease.

**Table S4.** **Characteristics of Patients Stratified by Circulating Targets Expression Levels Evaluated by ELISA.**

| Characteristics ^a^ | ABO^low^  N=30 | ABO^high^  N=30 | Pvalue | APOA5^low^  N=30 | APOA5^high^  N=30 | Pvalue | FN1^low^  N=30 | FN1^high^  N=30 | Pvalue | ROR1^low^  N=30 | ROR1^high^  N=30 | Pvalue |
| --- | --- | --- | --- | --- | --- | --- | --- | --- | --- | --- | --- | --- |
| Demographics and history | | | | | | | | | | | | |
| Age, median (IQR) | 66 (12.00) | 65.5 (12.00) | 0.773 | 66.5 (10.00) | 65 (13.00) | 0.310 | 66.5 (10.00) | 65.5 (13.00) | 0.420 | 66 (10.00) | 66 (12.00) | 0.657 |
| Male, n (%) | 21 (70.0%) | 19 (63.3%) | 0.584 | 19 (63.3%) | 21 (70.0%) | 0.584 | 19 (6.3.3%) | 21 (70.0%) | 0.584 | 22 (73.3%) | 18 (60.0%) | 0.273 |
| BMI>28 (kg/m^2^), n (%) | 3 (10.0%) | 6 (20.0%) | 0.472 | 5 (16.7%) | 4 (13.3%) | 1.000 | 5 (16.7%) | 4 (13.3%) | 1.000 | 2 (6.7%) | 7 (23.3%) | 0.145 |
| Hypertension, n (%) | 10 (33.3%) | 16 (53.3%) | 0.118 | 11 (36.7%) | 15 (50.0%) | 0.297 | 12 (40.0%) | 14 (46.7%) | 0.602 | 11 (36.7%) | 15 (50.0%) | 0.297 |
| Diabetes, n (%) | 17 (56.7%) | 15 (50.0%) | 0.605 | 17 (56.7%) | 16 (50.0%) | 0.605 | 19 (63.3%) | 13 (43.3%) | 0.121 | 13 (43.3%) | 19 (63.3%) | 0.121 |
| CVD, n (%) | 10 (33.3%) | 9 (30.0%) | 0.781 | 9 (30.0%) | 10 (33.3%) | 0.781 | 10 (33.3%) | 9 (30.0%) | 0.781 | 6 (20.0%) | 13 (43.4%) | 0.052 |
| Stroke, n (%) | 2 (6.7%) | 1 (3.3%) | 1.000 | 1 (3.3%) | 2 (6.7%) | 1.000 | 2 (6.7%) | 1 (3.3%) | 1.000 | 0 (0.0%) | 3 (10.0%) | 0.237 |
| Non-O blood type, n (%) | 8 (26.7%) | 6 (20.0%) | 0.542 | 11 (36.7%) | 3 (10.0%) | **0.015** | 12 (40.0%) | 2 (6.7%) | **0.002** | 5 (16.7%) | 9 (30.0%) | 0.222 |

BMI, Body Mass Index; CVD, coronary heart disease.

^a^ Divided into high and low expression groups based on the median peripheral blood expression levels of these targets. Propensity score matching (PSM) was not performed to balance covariates, due to the limited sample size.

**Table S5.** **Characteristics of Patients Stratified by IHC Score Levels.**

| Characteristics ^a^ | ABO^low^  N=48 | ABO^high^  N=48 | Pvalue | APOA5^low^  N=28 | APOA5^high^  N=28 | Pvalue ^b^ | FN1^low^  N=48 | FN1^high^  N=48 | Pvalue | ROR1^low^  N=48 | ROR1^high^  N=48 | Pvalue |
| --- | --- | --- | --- | --- | --- | --- | --- | --- | --- | --- | --- | --- |
| Demographics and history | | | | | | | | | | | | |
| Age, mean (SD) | 64.188 (9.66) | 64.771 (8.85) | 0.758 | 65.036 (6.506) | 65.000 (6.577) | 0.984 | 64.229 (10.31) | 64.729 (8.09) | 0.792 | 63.354 (10.58) | 65.604 (7.57) | 0.234 |
| Male, n (%) | 17 (35.4%) | 23 (47.9%) | 0.301 | 11 (39.3%) | 14 (50.0%) | 0.591 | 20 (41.7%) | 20 (41.7%) | 1.000 | 16 (33.3%) | 24 (50.0%) | 0.147 |
| Tumor Stage (%) |  |  | 0.479 |  |  | 0.297 |  |  | 0.378 |  |  | 0.402 |
| Stage I | 28 (58.3%) | 24 (50.0%) |  | 16 (57.1%) | 16 (57.1%) |  | 29 (60.4%) | 23 (47.9%) |  | 25 (52.1%) | 27 (56.3%) |  |
| Stage II | 17 (35.4%) | 18 (37.5%) |  | 11 (39.3%) | 7 (25.0%) |  | 16 (33.3%) | 19 (39.6%) |  | 20 (41.7%) | 15 (31.3%) |  |
| Stage III | 3 (6.3%) | 4 (8.3%) |  | 1 (3.6%) | 4 (14.3%) |  | 3 (6.3%) | 4 (8.4%) |  | 3 (6.3%) | 4 (8.3%) |  |
| Stage IV | 0 (0.0%) | 2 (4.2%) |  | 0 (0.0%) | 1 (3.6%) |  | 0 (0.0%) | 2 (4.2%) |  | 0 (0.0%) | 2 (4.2%) |  |
| CA199 > 37 U/mL (%) | 43 (89.6%) | 36 (75.0%) | 0.109 | 23 (82.1%) | 22 (78.6%) | 1.000 | 41 (85.4%) | 38 (79.2%) | 0.592 | 43 (89.6%) | 36 (75.0%) | 0.109 |

PSM, Propensity score matching; CA199, Carbohydrate antigen 199.

^a^ Grouping was based on the IHC score ratings of these targets, with negative and weak defined as low expression, and moderate and strong defined as high expression.

^b^ In the APOA5^low^ and APOA5^high^ group, PSM was performed to adjust the imbalanced variable age, using the greedy nearest-neighbor matching algorithm (1:1 matching ratio, caliper width of 0.2 SDs).
